# Markedly heterogeneous COVID-19 testing plans among US colleges and universities

**DOI:** 10.1101/2020.08.09.20171223

**Authors:** A. Sina Booeshaghi, Fayth Tan, Benjamin Renton, Zackary Berger, Lior Pachter

## Abstract

As the COVID-19 pandemic worsens in the United States [1], colleges that have invited students back for the fall are finalizing mitigation plans to lessen the spread of SARS-CoV-2. Even though students have largely been away from campuses over the summer, several outbreaks associated with colleges have already occurred [2], foreshadowing the scale of infection that could result from hundreds of thousands of students returning to college towns and cities. While many institutions have released return-to-campus plans designed to reduce viral spread and to rapidly identify outbreaks should they occur, in many cases communications by college administrators have been opaque. To contribute to an evaluation of university preparedness for the COVID-19 pandemic, we assessed a crucial element: COVID-19 on-campus testing. We examined testing plans at more than 500 colleges and universities throughout the US, and collated statistics, as well as narratives from publicly facing websites. We discovered a highly variable and muddled state of COVID-19 testing plans among US institutions of higher education that has been shaped by discrepancies between scientific studies and federal guidelines. We highlight cases of divergence between university testing plans and public health best practices, as well as potential bioethical issues.

## Heterogeneity in test strategy

To survey the return-to-campus testing strategies across the US, we curated a database of testing plans for 1226 institutes of higher education in the US, based on a compilation published in the Chronicle [3]. Examining the reopening plans of over 500 schools, we found, as of August 7, 2020, highly variable strategies all over the country, covering the full spectrum from available and regular testing for students, staff and faculty, to no testing at all. Our findings reveal that 54% of universities are performing or facilitating some form of COVID-19 testing. Only 27% of colleges are performing initial re-entry testing at least for undergraduates, while an even smaller percentage, 20%, plan to test their communities regularly to some extent.

Our survey also showed a distinct geographical bias in testing. We found, for example, that universities in the Northeast plan to offer extensive testing. Many schools are able to do this thanks to testing capacity developed by one institute: 11 schools in Massachusetts, three in New York, two in Maine, one in Rhode Island, and one in Connecticut are all contracting with the Broad Institute to perform their testing. Of concern is the fact that in the five states (MA, NY, PA, CA, TX) with the most universities and for which we had extensive data, we find that states with the highest COVID-19 test positivity rate plan for the least amount of initial testing of students returning to campus (**Figure 1a**). The concern stems from the fact that institutions that are implementing initial testing are identifying infectious individuals who in the absence of quarantine could spark outbreaks [4].

**Figure 1:**
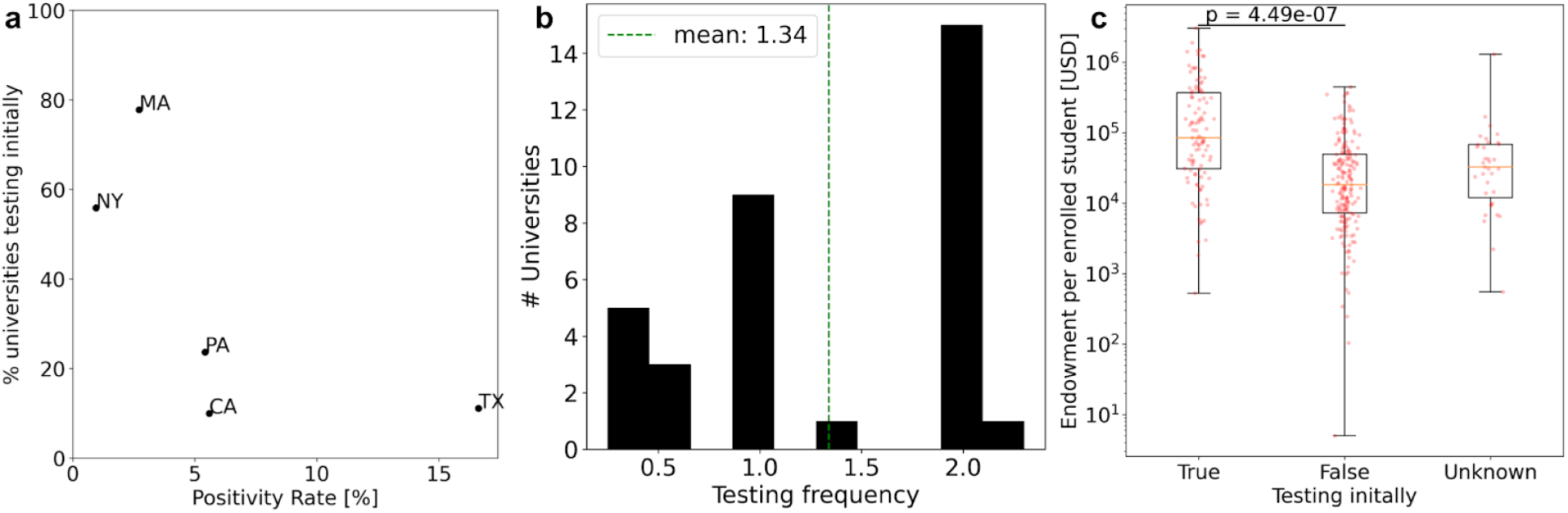
(a) The weekly-averaged positivity rate in New York, Massachusetts, Pennsylvania, California and Texas versus the percent of universities performing initial testing. (b) The reported frequency of tests per week at schools that plan on performing recurrent screening of their communities. (c) The initial (return to campus) testing plans for each university compared to the per-enrolled-student endowment. A two-sided *t-test* was performed between universities that are and are not testing initially upon return to campus.

In the absence of a collective, nationwide testing strategy specifically focused on universities and colleges, and with the market price for tests effectively set by the Centers for Medicare and Medicaid Services [5], there is immense pressure on institutions to maximize the utility of every test [6]. Either in place of, or in conjunction with re-entry testing, many schools urge students to undergo 14 day quarantines, or require the submission of PCR test results taken from a certain time period prior to their return. The submission period for PCR tests varies greatly— as short as 10 days before arrival in the case of Syracuse University in New York [7], and as long as 30 days for Simpson University in California [8]. At USC, students are recommended to not retest if they have received a positive test result within the 90 days prior to returning to campus [9] despite the conflicting data on the duration of neutralizing antibodies after COVID-19 infection [10]. Meanwhile, some universities are exploring additional ways of monitoring their community using methods for which data on effectiveness is still scarce. The University of South Florida is planning to conduct pooled testing of environmental DNA [11], and Michigan Tech is planning to test wastewater [12].

## The scientific consensus on testing

The variability of testing strategies might lead one to conclude that there is uncertainty in the science of best practices for safe reopening of colleges. However, contrary to that perception, recent research has converged on the need for easily accessible, frequent testing for colleges to reopen safely. Though high-sensitivity PCR tests are viewed as the gold standard in terms of test sensitivity, recent work suggests that the frequency and turnaround times for COVID-19 tests are more important than the sensitivity of the tests administered [13,14]. This is corroborated by research that supports the frequent testing approach for college campuses specifically, finding that testing college communities once every two days would be sufficient to contain outbreaks on campuses [15]. However, not all colleges are planning to regularly screen their communities, and even those who do display high variability in the frequency with which they plan to test (**Figure 1b**). One reason might be that testing students comes with a significant cost. In its decision to go fully online, the Chancellor of the California State University system, Timothy White, cited the fact that testing half the system’s student population weekly would cost $25 million, a financial burden the system could not bear [16]. However, while cost is prohibitive for some universities, others such as the University of Georgia have invested $1.2 million dollars to procure 24,000 tests, enabling 300 members of the community to be tested per day, every day, until the fall term ends at Thanksgiving [17]. Another issue is government policy and recommendations. The current CDC messaging for college administrators presents guidance on mitigative measures, such as cleaning, communication and contact tracing [18]. However, despite an abundance of literature on the importance of regular and frequent testing, testing for COVID-19 is presented as an “interim consideration” [19]. The guidelines amount to a lack-of-evidence non-recommendation [20], stating that because it is “unknown if entry testing in institutes of higher education provides any additional reduction in person-to-person transmission of the virus”, the CDC “does not recommend entry testing of all returning students, faculty, and staff” [9]. Now there is growing evidence that universities are using lax federal guidelines to justify the lack of testing on their campuses. For example, the University of North Carolina at Chapel Hill explicitly references CDC guidelines in its decision not to test returning students, placing the responsibility on individuals to take preventative measures, as testing everyone would create “a false sense of security” [21]. The University registered 175 cases of COVID-19 on campus as of August 5th 2020, as its undergraduates returned to dorms that same week [22]. At some universities where there are no plans for COVID-19 testing or screening, administrators have given up all pretense of a rational defense of their policies. At Pepperdine university the lack of screening plans is excused by claiming that “Mass screening has limited efficacy (as it only establishes a result in a moment in time)” [23].

## Discrepancies in testing plans and equity

Distributive justice, which concerns the socially just allocation of resources, is a key bioethical principle which the COVID-19 pandemic in the US has shown to be routinely violated in the healthcare sector [24]. Many of the equity problems in healthcare have now spilled over to higher education in the context of the COVID-19 pandemic. We find that disparities exist between universities based on their resources: more private universities plan on testing than public (37% vs 16%), and many more of the US colleges and universities that are ranked in the top 50 by US News World Report [25] plan on testing, compared to less highly ranked institutions (96% vs 49%). Furthemore, universities with higher endowments are more likely to plan for testing than universities with smaller endowments (**Figure 1c)**. Universities that will not provide testing, perhaps due to cost or other considerations, are in some cases offering bizarre rationales. At Newberry College in South Carolina, authorities claim that “testing is not required because the tests available at this time cannot provide assurance that someone will not become sick after the test is performed.” [26]

Yet disparities within universities are just as important and in many cases even more extreme. We found that public-facing websites generally lacked information on whether staff or contract workers were offered testing, consistent with evidence of numerous disparities in their treatment by universities during the COVID-19 pandemic [27]. Such workers, who are typically more racially and ethnically diverse than students or faculty [28], are also at higher risk of serious complications from COVID-19 [29]. Universities thus seem to be reinforcing societal disparities with their COVID-19 testing policies. Moreover, at some schools, students and staff are seemingly being prioritized as study subjects, instead of a focus on their safety as they are slated to return to campus. For example, Pennsylvania State University presented the return of 40,000 students as a “research opportunity”, a chance to study pandemic outcomes in real time. However, with uncertainties around reopening, some residents of the small college town community of State College borough are concerned that this research opportunity might turn into a public health crisis [30].

Only with urgent action on the part of Federal and state authorities, working in concert with university administrators, will the reopening of universities in the fall avoid a public health crisis that worsens the national pandemic. Such action should be guided by transparency, universal access to testing on the part of all who participate in university life, and decision making based on science and sound ethical principles.

## Data Availability

The database of college and university testing plans is available as a Google Sheet. The code to generate all of the figures and results in the manuscript can be found on Github.

https://docs.google.com/spreadsheets/d/10I8bVkLzvrmXJsb5N-8JSFpWw5vBwDKYzyOVAI4viKo/edit#gid=1514440859

https://github.com/pachterlab/BTRBP_2020

## Acknowledgments

We thank the numerous anonymous contributors to the testing database, who helped collate information about testing plans of colleges and universities throughout the United States.

Thanks to Carl Bergstrom for facilitating the collaboration that led to this work via introductions of several of the authors, and to Ingileif Hallgrimsdottir and Laura Luebbert for comments that improved the manuscript. We thank the COVID-19 committees at Caltech for bringing to our attention many of the issues facing colleges and universities planning to reopen for the 2020 Fall Semester.

## Author contributions

ASB and LP started compiling the testing database; FT filled in the majority of entries. FT drafted the initial manuscript; ASB, FT, ZB and LP wrote the manuscript. ASB performed the analysis of the testing database and made the figures. BR analyzed college endowments and their relationship to testing plans. ASB, FT, BR and LP contributed references.

### Data and software availability

The database of college and university testing plans is located here. Code to generate all of the figures and results in the manuscript can be found here.

### Funding declaration

None.

### Conflict of interest declaration

The authors declare no conflicts of interest.

## Notes

### Competing Interest Statement

The authors have declared no competing interest.

### Funding Statement

No external funding was received in support of this project.

### Author Declarations

This project did not require IRB approval.

